# A Community-Based Participatory Research to Assess the Feasibility of Ayurveda Intervention in Patients with Mild-to-Moderate COVID-19

**DOI:** 10.1101/2021.01.20.21250198

**Authors:** Vishwesh Kulkarni, Neha Sharma, Dipa Modi, Abnimanyu Kumar, Jaydeep Joshi, Nagamani Krishnamurthy

## Abstract

Innovative strategies are required to manage COVID-19 in the communities. Back to Roots was a collaborative, community-based pilot intervention project in the British Asian community. To assess the efficacy and safety of Ayurveda intervention in relieving symptoms of mild-to-moderate COVID-19, a community based participatory research framework was used. Twenty-eight patients were enrolled with confirmed COVID-19 clinical stages of mild-to-moderate COVID-19, symptomatic, and between 20 to 70□years of age. Routine management was followed by all patients managing at home, additionally patents taking Ayurveda intervention for 14 consecutive days. The efficacy and safety of Ayurveda intervention were evaluated. There were suggestions of Ayurveda’s advantage in improved symptoms relief, clinical recovery in 7 days. However, a control group was not included but data triangulations from separate usual care found the difference statistically significant. Ayurveda intervention may potentially have a beneficial effect on patients with COVID-19, especially for those with mild to moderate symptoms. A further definitive large-scale clinical trial is necessary.

**ClinicalTrials.gov Identifier:** **NCT04716647**

## Introduction

Covid-19 outbreak became a global health crisis that is unparalleled in modern history with most locked down most of the world for months. Despite the contact and rigorous efforts from world scientific experts, there is currently no proven prophylaxis for those who have been exposed to Covid-19 nor treatment for those who go on to develop COVID-19. The World Health Organisation (WHO) has recommended symptomatic management to help relieve symptoms, for severe cases, treatment should include care to support vital organ functions and no specific antiviral treatment for COVID-19^1^. The absence of specific drug treatment is a critical gap in the knowledge of the COVID-19 pandemic.

The Ayurveda, Indian Traditional system of medicine, recommends a variety of herbal formulations for the diseases affecting the respiratory tract.^2-5^ Ayurvedic medicinal plants *Withania somnifera* (Ashwagandha)^6,7^, *Tinospora cordifolia* (Giloy)^8,9,^ and *Ocimum sanctum* (Tulsi)^10,11^ has been highly effective for the treatment of respiratory diseases and fever. A recent in-silico study highlights the significance of phytochemicals identified in *Tinospora cordifolia* in controlling SARS-CoV-2 replication.^12^ *Tinospora cordifolia* is already well known to be responsible for various antiviral properties^13-15^ and the treatment of COVID-19 asymptomatic patients.^16^

Another therapeutic agent against COVID-19 has been found in the plant *Withania somnifera* using molecular docking approaches.^17-19^ Two research groups identified WFA Withanolides, which bind to the viral S-protein receptor binding domain, thereby blocking or reducing interactions with host ACE2 receptor.^17,18^ Similarly third research group reported Withanone, a different form of withanolide, interact with the main protease of SARS-CoV-2.^19^ Previously, *Ocimum sanctum*, has been proven effective in many in-vitro, animal, and human studies including antimicrobial, anti-inflammatory, and immunomodulatory effects.^20-23^ Recent Molecular docking study also found *Ocimum sanctum* to have high binding efficacy against SARS-CoV-2 targets involved in viral attachment and replication.^24^

Altogether, *Withania somnifera* (Ashwagandha), *Tinospora cordifolia* (Giloy), *Ocimum sanctum* (Tulsi) has been reviewed as a remedial option against COVID-19.^25^ Molecular docking analysis identified six phytochemicals from Ashwagandha, Giloy, and Tulsi with potential inhibition and high affinities towards SARS-CoV-2 predicted to restrain the action of SARS-CoV-2 thereby obstructing further translation of viral protein that assists in damaging the vital organs of the host.

Ayurveda combination of Giloy, Ashwagandha, and Tulsi had been widely used by people and recommended by Ayurveda practitioners for suspected and confirmed COVID-19 cases for relieving the symptoms and preventing the escalation of severity and complications of mild-to-moderate COVID-19. To further evaluate the efficacy and safety of the combination, present study was undertaken.

## Methods

Symptomatic relief, prevention of severe infection or complications, and easy availability of care for everyone are some of the priorities of the COVID-19 pandemic. Focusing on the priorities, Community based project Back to Roots was set up in Leicester, UK during the early outbreak of COVID-19, which also aimed at improving the health literacy of patients with COVID-19 as well as at reducing the risk of severe or adverse side effects due to ill-advised self-medication and overmedication with over the counter Ayurvedic or herbal interventions. The aims of the Back to Roots community project were to support a safe practice of easily available remedies that pose no risk; to improve the self-efficiency in the community participants and to evaluate that the participants who succeed in improving symptoms can reduce the incidences concerning the severity of infections and COVID-19 related complications. We hypothesise that an interactive community based participatory program may increase patients’ compliance with standard recommendations and decrease self-medication or over medication of herbal supplements. People who wanted to use Ayurveda, encouraging them to do so under the supervision of qualified practitioners and with COVID-19, participants have safe recovery.

Participants who enrolled for the community support program ‘Back to Roots’ were registered with their clinical symptoms and medical history, further reviews by experienced Ayurveda practitioners who could provide them with standard recommendations and best suited supplements to manage the symptoms as per individualisation, monitor the variations in the vital signs and symptoms of the participants.

Pregnant and lactating women were not advised any herbal supplements. The exclusion criterion was extended to cover individuals with (1) any of the known COVID-19 complications and emergency conditions which may require shift/admission in intensive care unit, (2) chronic, severe and uncontrolled comorbid medical conditions, (3) active status on immunosuppressants, steroids, or chemotherapy, (4) cancer or a history of peptic ulcer or bleeding or chronic pulmonary diseases (COPD) and (5) Ayurveda practitioners refusing based on individual medical condition. In excluded condition, participants were supported only through food and diet recommendation, regular observation and standard recommendations following guidelines of National Health Services.

### Experimental design

In the community support project, participants were recommended Ayurveda based supplements as per the individualisation. From the support project, 28 patients, aged 20-70 years with a diagnosis of mild-to-moderate COVID-19 infection who had used Ashwagandha, Giloy and Tusli combinations were further analysed retrospectively by research team.

### CBPR PARTNERSHIP

CBPR partnership was set up through:

1. Special interactive online community groups for keeping in contact with recruited participants.
2. Dissemination of the project information to the public -- flyers distributed through pharmacies, doctors, charities, etc. On the basis of previous experience, word of mouth among patients who were enrolled in Aarogyam projects were expected to be the most effective recruitment strategy.

Patients were enrolled on a voluntary basis, signed informed consent through the app, filled daily vitals and symptoms score. The vital data and changes in the symptoms together with the self-efficiency score were monitored through a 14-day period. Patients were followed up for 2-weeks post-intervention for any Post COVID symptoms.

### Ayurveda Intervention

Ashwagandha, Giloy, and Tulsi were given in tablet form for oral administration. Dosage was used in a common range (Ashwagandha: Doses range from 250 mg to 5 g; Giloy: range from 500mg to 1g; Tulsi: 500mg-1g) Dosage were altered based on age, weight, and severity of symptoms. Patients were given diet recommendations and were followed regularly.

### PHASES OF CBPR

The scientific method of the Back to Roots study includes three phases.

- Phase I started in September 2020. During the first phase, we created a community forum and community-based support group. A *Community Advisory Board* (CAB) was set up which reviewed the need assessment and a summary of the project in plain language and instructions to enrol for the second phase.
- Phase II started on October 9, 2020. Eligible patients were invited who were willing to participate in the project. Their initial assessment was taken over the phone by an Ayurveda consultant, who decided the Ayurveda combination for all patients. From the day of enrolment, all participants were asked to score symptoms severity regularly, each day for 14 days. During this phase, we also encouraged Back to Roots participants to meet online, exchange the information, experience, and on the strategy to comply with the recommendations.
- In Phase III, Patients that met the eligibility criteria and were prescribed Ayurveda intervention decided for the study, were further analysed by academic partners (research) team retrospectively.

### Data Triangulation

In concurrent forms of data collection was used to transform the usual care group data for comparison with the intervention group as a control. A total of 19 mild to moderate Covid-19 patients receiving usual care at home were analysed using mixed methods. Not only are the control group data were retrospectively collected during the same timeframe and independent of each other, but they were also collected from different community support organizations who were supporting families during the pandemic. Control Group participants were not enrolled in the study. Participant’s information from separate community initiatives was taken retrospectively for the comparison purpose only.

### Ethics

The Ayurveda intervention chosen by us does not interfere with the standard management of COVID-19. The participants themselves shared the information on the treatment received by them.

Under the CBPR framework, each community participants were made aware of research and evaluation at the end of the program with anonymised data only. Informed consents were taken during the first assessment call to share anonymised efficacy and safety data for evaluation. The major principles of ethics while conducting a CBPR study was followed rigorously. The following aspects of ethics which usually create a challenge was taken care of:

- Mutual respect – Community Advisory Board (CAB) meetings were conducted with the understanding that everyone should listen to a member’s opinion with respect without judging the same.
- Democratic participation – All members of the CAB were given equal opportunity to propose their ideas towards the intervention.
- Equality and inclusion – Members were selected with no bias of gender, ethnicity, age, class, education, (dis)ability, faith or sexual orientation.
- Data management – Data were monitored and managed by the community organisation’s Data Safety and Monitoring Board following General Data Protection Regulation (GDPR).
- Anonymity-Data collected during the community program was anonymised removing personal identifiers, both direct and indirect, that may lead to an individual being identified. ICO’s Code of conduction on Anonymisation was followed. ^31^
- Privacy – Personal data of participants were protected and safeguarded as per the UK law and regulation.
- Confidentiality – An informed consent was taken by the participants before they voluntarily participated in the community project.
- Interpretation of results – Data was interpreted without mentioning the personal details of any of the participants. Participants cannot be identified by any reports or data shared with university.
- Ownership – The primary data was stored in the organisation’s archive and only anonymised secondary data was provided for analysis, interpretation and publication. Secondary data from the study was transferred using AiM COVID App designed by the University of Warwick and used only for evaluation purposes.

### Endpoint

The primary efficacy endpoint was time to clinical recovery (mean days for clinical recovery, Day of enrolment to the day of clinical recovery). The secondary outcome was the rate of patients with negative SARS-CoV-2 on nasal or throat swab. Due to logistical issues at the time of study implementation, virological measurements were limited, including SARS-CoV-2 polymerase chain reaction tests on day 7 only.

### Data Collection and Analysis

Data were collected from patients and Ayurveda physicians using electronic transfer and stored in organization;s cental facility with limited access. The “AiM COVID-19” app available on the “ClickMedix” platform was used to transfer the secondary data to research analysis team. All statistical processing was performed using SPSS software. The data were individually analyzed for central tendencies (mean, median), range, standard error, standard deviation, and 95% confidence intervals for each intervention arm in each of the groups in the study. Though ANOVA was used for most of the measures, standard parametric (Student t-test) and non-parametric statistical tests (Mann Whitney statistic, Kruskal Wallis test) were used depending upon the normality of the data. Chi-Square was used to test the proportion of patients.

## Results

In all, 42 patients were screened as per inclusion criteria; 28 patients who were eligible for inclusion and focus Ayurveda combinations were analysed.

All 28 included patients completed the intervention and post-intervention for 2-weeks. The characteristics of the patients are shown in Table 1. Most patients (78.5%) started Ayurveda supplements intervention within 3 days after symptom onset. The mean age of the patients was 45.1 years, and 53.6% of all the included patients were men.

**Table 1:** Baseline characteristics of study participants.

| **Characteristics** | **Ayurveda supported group**<br>**(n=28)** | **Usual Care Group**<br>**(n=19)** |
| --- | --- | --- |
| **Age**, years; mean (SD) | 45.1 (11.8) | 47.7 (8.82) |
| **Gender**, F n (%) | 13 (46.4) | 12 (63.1) |
| **Ethnicity** |  |  |
| Indian | 23 (82.1) | 16 (84.2) |
| Pakistani | 2 (7.1) | 2 (10.5) |
| Bangladeshi | 3 (10.7) | 1 (15.7) |
| **Health care worker n (%)** | 5 (17.8) | 2 (10.5) |
| **Comorbid health condition** |  |  |
| Diabetes | 5 (17.8) | 4 (21.1) |
| Hypertension | 4 (14.2) | 6 (31.5) |
| Asthma | 3 (10.7) | 5 (26.3) |
| None | 20 (76.92) | 8 (42.1) |
| **Days of Symptoms onset to enrolment; Mean (SD)** | 3.78 (0.9) | 6.6 (1.1) |

None of the enrolled patients had chronic lung disease, chronic kidney disease, autoimmune disease, or immunodeficiency disease. A significantly higher number of patients had asthma, and hypertension as a comorbid condition in the usual care group; the Ayurveda group had a significantly higher number of participants without comorbid health conditions compared to the control group (difference 34.82%; p<0.01). Days of symptoms onset to enrolment in the support group were significantly more in the control group compared to Ayurveda intervention (mean difference, 2.820: P < 0.0001) (Table 1)

Sixteen patients (57.1%) patients in the Ayurveda group, experienced fever and cough. None of the patients complained of dyspnea, diarrhea, palpitation, or nausea, vomiting, abdominal pain, or chest pain in the Intervention group. (Table 2)

**Table 2:**
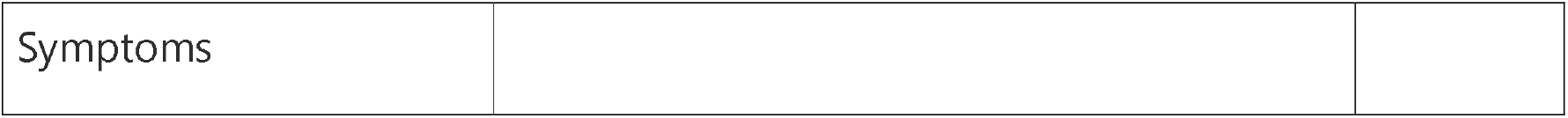
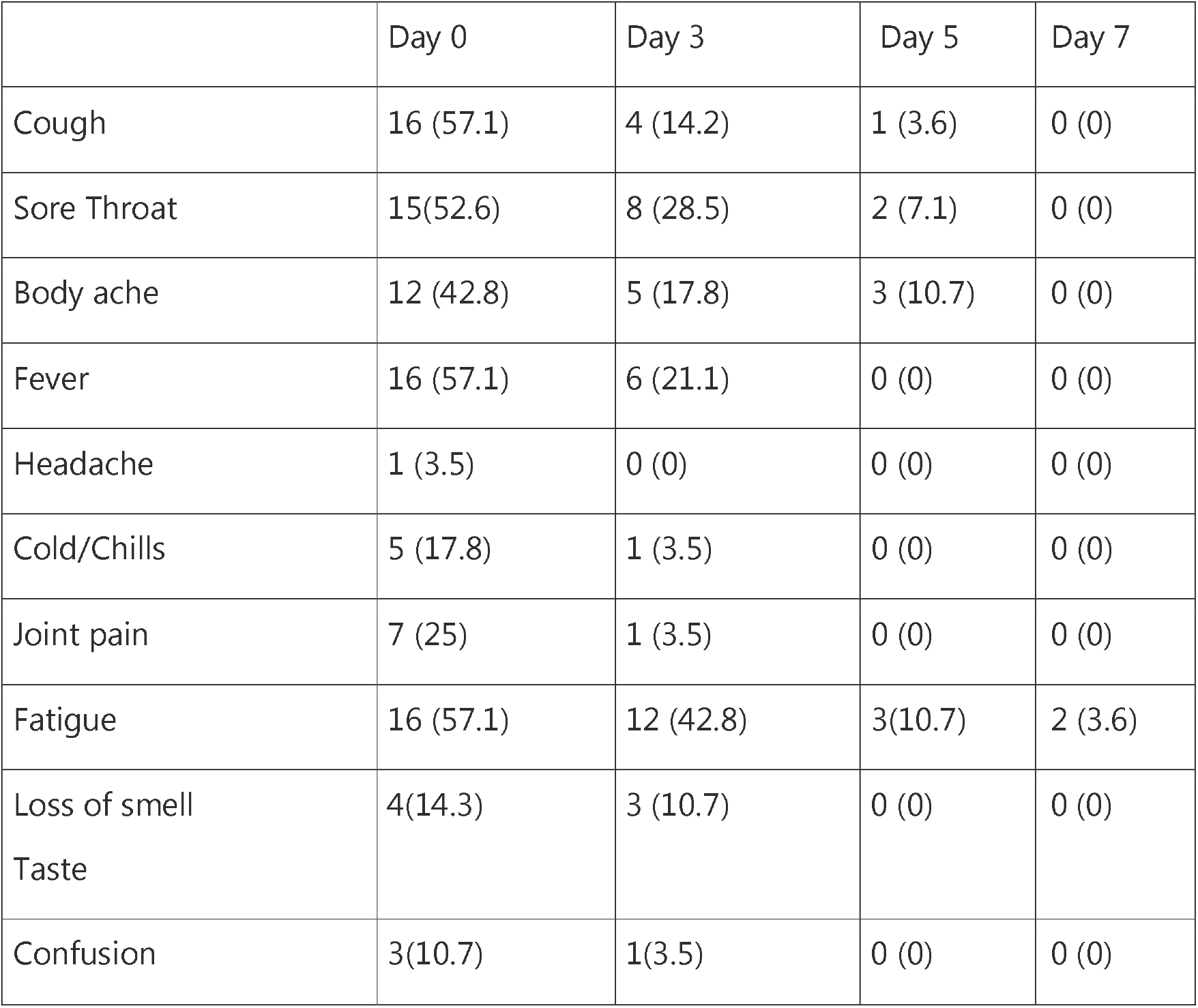
Symptoms recovery in the Intervention group.

### Efficacy Outcomes

The average time to recover was 4.85 (SD 1.8) days in the Ayurveda group, and 13.5 (SD 6.4) in the control group, with the highly statistical difference between them (P < 0.0001) (Table3; Figure 2). Full recovery on day 3 was seen in 10 (35.7%) patients, on day 5, 22 (78.5%), and on day 7, 26 (92.8%) patients recovered fully. Fatigue was the only symptom reported on day 7 in two patients. None of the patients reported Long Covid symptoms at the 2-weeks post-intervention mark.

**Table 3:**
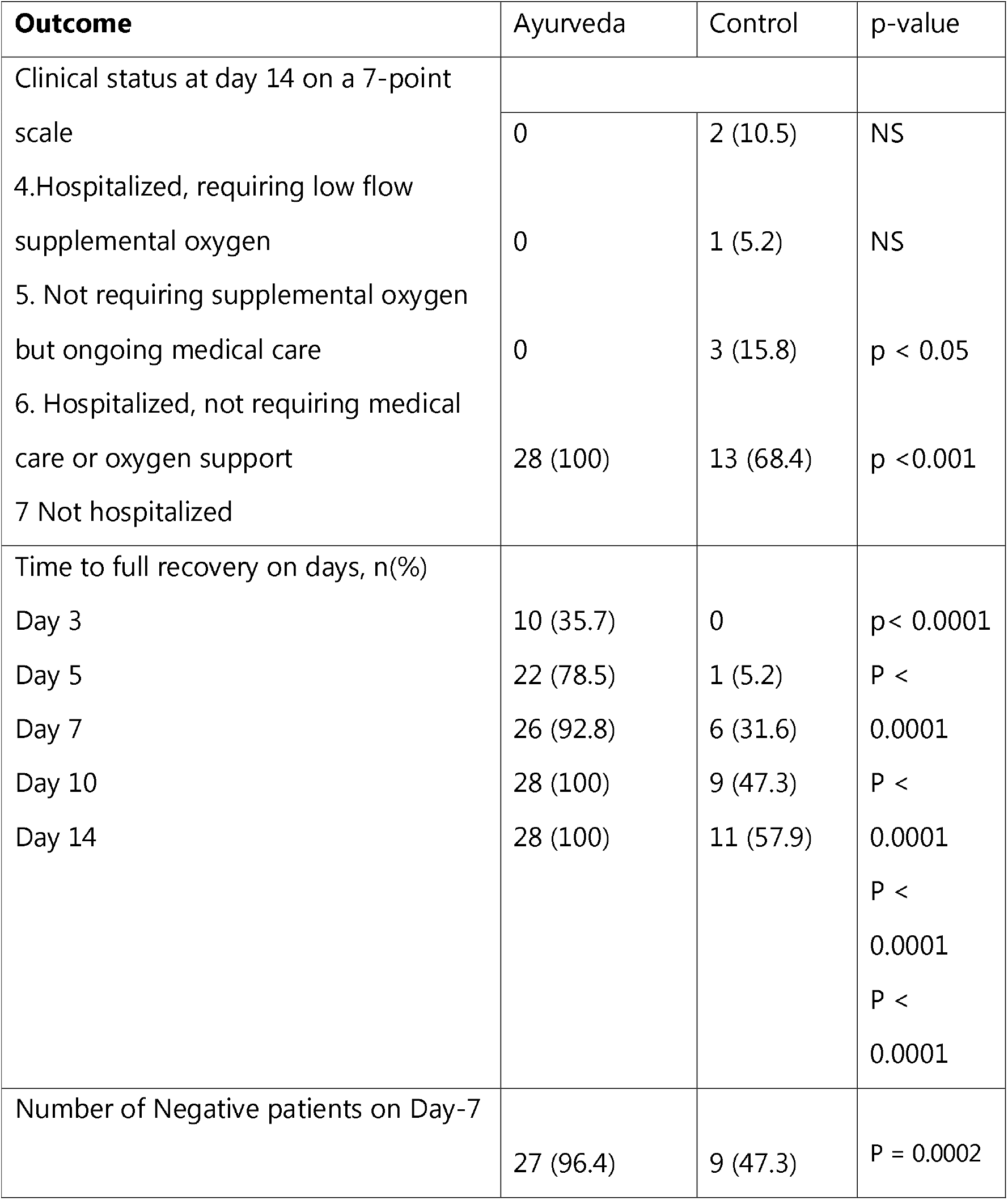
Clinical Outcome.

**Figure 2:**
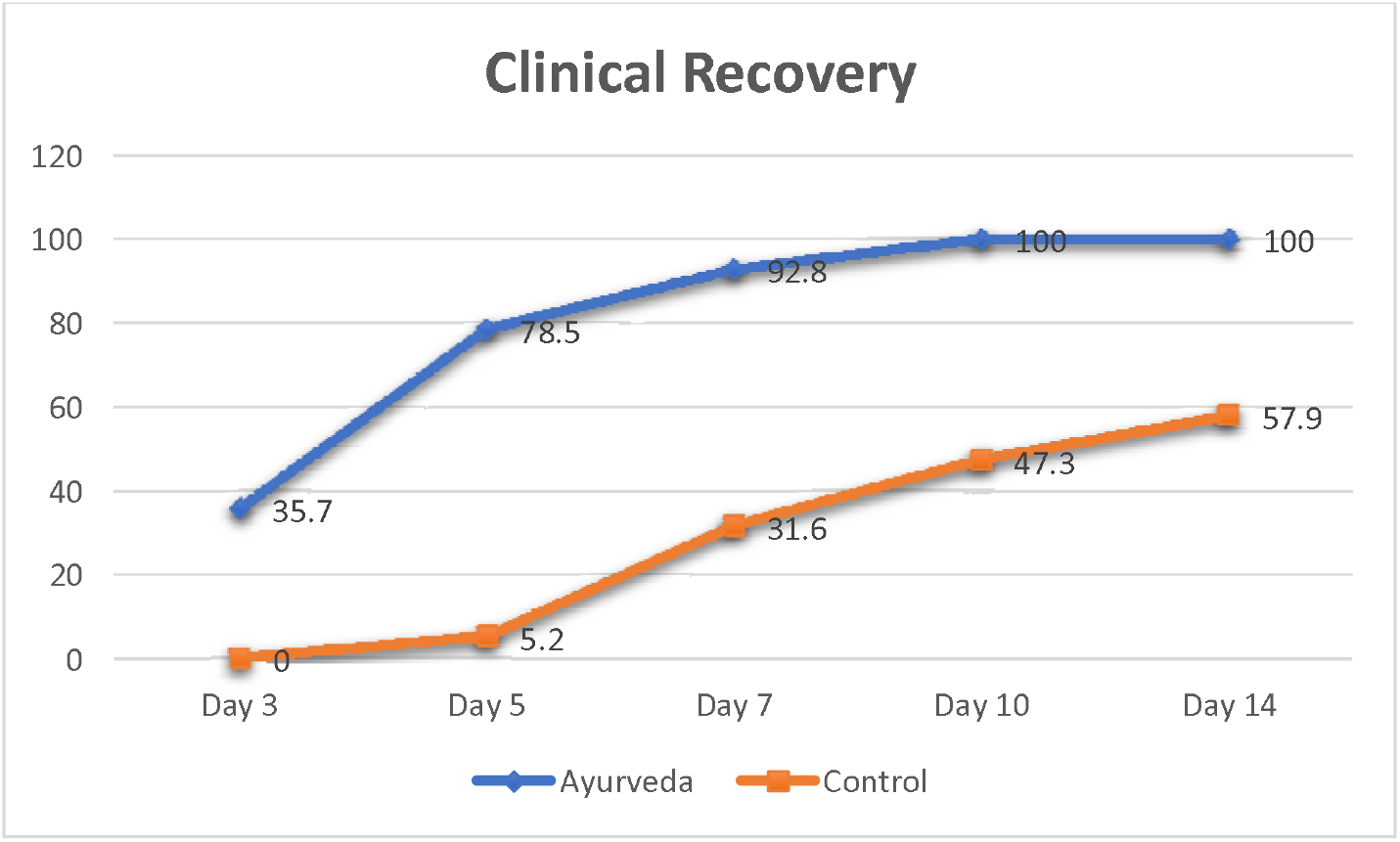
Clinical Recover over the 14-days period.

After 7 days of treatment, the positive-to-negative conversion rates of SARS-CoV-2 nucleic acid in the pharyngeal swab in the Intervention group, and the control group were 96.4% (27/28), and 47.3% (9/19), respectively, and did present a statistical difference among the groups (p <0.001).

On a 7-point ordinal scale of COVID-19, no participants were hospitalized for any medical or oxygen requirements. None developed breathing difficulty during the intervention period or post-intervention. In the usual care, two patients (10.5%) required low flow supplemental oxygen having underlying asthma (5.2%) and diabetes (5.2%). One patient was taken to the hospital with diabetes and hypertension. Three patients (15.5%) were required to be taken into the hospital with breathing concerns but not required oxygen support or further medical assistance.

### Potential biases and limits

For the Back to Roots Project, its inclusion of a greater proportion of patients who had undergone Ayurvedic treatments for symptomatic relief in the past may influence the information collected in that project. Another limitation was the lack of a control group.

## Discussion

For ethical reasons and because results are significant and evident in the present context, we decided to share our findings, given the urgent need for effective management and an overwhelmed health care setting against SARS-CoV-2.

We show here that Ayurveda self-management is efficient in clearing viral nasopharyngeal carriage of SARS-CoV-2 in COVID-19 patients in only seven days, in most patients. A significant difference was observed between Ayurveda managed patients and usual care starting even from day 3 post-intervention. These results could be of great importance as a consortium study of mild-to-moderate COVID-19 patients reported a mean disease duration of 11.5□±□5.7 days.^26^ Another Chinese study of COVID-19 patients observed an average time to recovery of 10.63±1.93 days for mild to moderate patients and 18.70±2.50 for severe patients.^27^ Two more single-center Chinese studies (127 and 225 recovered patients) found a mean recovery time or median time of 20 and 21 days, respectively.^28,29^ Findings are similar in the usual care compared in our study with an average time to recover was 13.5 (SD 6.4) with none having time to recovery according to patient age or sex.

Ayurveda intervention starting at early onset (3.78 average days) may have influenced the early recovery on average 4.85 (SD 1.8) days. Ayurveda findings are similar to recently published results of a study demonstrating that the Ayurveda combination appeared to accelerate recovery of patients hospitalized for COVID 19 infection, in terms of reduction of symptoms and duration of hospital stay.^30^

Our preliminary results also suggest a similar effect of the combination of Ayurveda to prevent severe respiratory tract infections when administrated to patients suffering from viral infection. Further studies on this combination are needed, since such combination may both act as an antiviral therapy against SARS-CoV-2 and prevent bacterial super-infections.

Such results could be promising and open the possibility of an international strategy to decision-makers to fight this emerging viral infection in real-time even if other strategies and research including vaccine development could be also effective. We, therefore, recommend that mild to moderate COVID-19 patients be self-managed with properly guided Ayurveda care in the community.

## Conclusions

A collaborative effort between patients and researchers to engage in research that benefits the community is a central tenet of CBPR. The aim of the present study was to improve the framework of CBPR with symptoms relief and prevention of severe infection of mild to moderate COVID-19. In particular, we report that an Ayurveda intervention, viz., Giloy, Ashwagandha, and Tulsi can significantly reduce symptoms severity and ultimately prevent the incidence of Covid-19 related complications.

## Data Availability

The datasets generated during and/or analysed during the current study are available from the corresponding author on reasonable request.

## Acknowledgments

We gratefully acknowledge the support of Dabur (India) for supplying the supplements for community initiative. We appreciate Hindu Swayamsevak Sangh (UK) and partner community support groups for their support in community outreach.

